# Spatial variability in reproduction number and doubling time across two waves of the COVID-19 pandemic in South Korea, February to July 2020

**DOI:** 10.1101/2020.07.21.20158923

**Authors:** Eunha Shim, Amna Tariq, Gerardo Chowell

## Abstract

**Objectives:** In South Korea, 13,745 cases of coronavirus disease (COVID-19) have been reported as of 19 July, 2020. To examine the spatiotemporal changes in the transmission potential, we present regional estimates of the doubling time and reproduction number (*R*_*t*_) of COVID-19 in the country.

**Methods:** Daily series of confirmed COVID-19 cases in the most affected regions were extracted from publicly available sources. We employed established mathematical and statistical methods to investigate the time-varying reproduction numbers of the COVID-19 in Korea and its doubling time, respectively.

**Results:** At the regional level, Seoul and Gyeonggi Province have experienced the first peak of COVID-19 in early March, followed by the second wave in early June, with *R*_*t*_ exceeding 3.0 and mean doubling time ranging from 3.6 to 10.1 days. As of 19 July, 2020, Gyeongbuk Province and Daegu are yet to experience a second wave of the disease, where the mean *R*_*t*_ reached 3.5-4.4 and doubling time ranging from 2.8 to 4.6 days during the first wave.

**Conclusions:** Our findings support the effectiveness of control measures against COVID-19 in Korea. However, the easing of the restrictions imposed by the government in May 2020 facilitated a second wave in the greater Seoul area.

**Highlights:**

- South Korea has experienced two spatially heterogenous waves of COVID-19.
- Seoul and Gyeonggi Province experienced two waves of COVID-19 in March and June.
- In the densely populated Seoul and nearby areas, reproduction numbers exceeded 3.0.
- The easing of the social distancing measures resulted in the second wave.

## Introduction

Since the first COVID-19 cases reported in Wuhan, Hubei Province, China, in December 2019, more than 24.7 million cases of coronavirus disease (COVID-19), including more than 830,000 related deaths, have been reported worldwide (WHO) as of August 30, 2020. In South Korea, the novel coronavirus was first diagnosed in a 36-year-old Chinese woman who entered the country on 20 January 2020. South Korea has since experienced two heterogenous waves of the disease with a total of 13,745 cases including 295 deaths as of 19 July 2020 (KCDC, 2020a).

During the early phase of the COVID-19 outbreak in South Korea, public health authorities primarily conducted strict contact tracing and isolation of confirmed cases as well as quarantined those suspected with the novel coronavirus (Covid-19 National Emergency Response Center et al., 2020). As the number of COVID-19 cases continued to increase, Korean public health authorities set the alert to the highest level (Level 4) on 23 February and mandated the population to report any symptoms related to COVID-19 for further screening and testing. In addition, the country rapidly adopted a “test, trace, isolate, and treat” strategy that has been deemed effective in stomping out localized outbreaks of the novel coronavirus (KCDC, 2020a). However, the total number of confirmed cases in South Korea spiked from 31 cases on 18 February to 433 on 22 February. According to the Korea Centers for Disease Control and Prevention (KCDC), this sudden jump was mainly attributed to a super-spreader (the 31^st^ case) who had participated in a religious gathering of attendees of the Shincheonji Church of Jesus in Daegu (KCDC, 2020a). These superspreading events occurred in the Daegu and Gyeongbuk provincial regions, leading to more than 5,210 secondary COVID-19 cases in Korea (KCDC, 2020a, Ryall, 2020). These events facilitated sustained transmission chains, with 38% of the cases in the country associated with the church cluster in Daegu (Shim et al., 2020b).

On 8 March, the KCDC announced that 79.4% of the total cases had epidemiological links, whereas the remaining 20.6% cases were either sporadic cases or under investigation (KCDC, 2020a). Case clusters started to accumulate from churches in the Seoul capital area, and on 17 March, 79 church attendees developed COVID-19 after attending a service at the River of Grace Community Church. Notwithstanding social distancing orders put forward by the government, some churches continued to conduct services, which led to new clusters of infections. For instance, the Manmin Central Church in Seoul was involved in one of the clusters, with 41 infections linked to a gathering in early March; SaengMyeongSu Church in Gyeonggi Province was another church cluster linked to 50 cases (Park, 2020).

As SARS-CoV-2 infection spread rapidly outside Korea, the number of imported cases started to increase, resulting in 476 imported (4.9%) cases out of 9,661 total cases as of 30 March. Consequently, as of 1 April, the KCDC implemented self-quarantine measures for travellers from Europe and the U.S.A (KCDC, 2020a). In addition, incoming travellers with symptoms but negative test results for coronavirus, as well as asymptomatic short-term visitors were ordered to follow a 2-week quarantine in the government facilities (KCDC, 2020a).

Such control measures undertaken by South Korea have been deemed successful in limiting the spread of the outbreak, without locking down entire cities (Normile, 2020). Therefore, after a sustained period of low incidence with fewer than 20 cases per day (16 April – 5 May), the government eased its strict nationwide social distancing guidelines on 6 May, with a phased reopening of schools starting mid-May, 2020. However, a new cluster linked to nightclubs in Itaewon emerged in central Seoul in early May, resulting in a resurgence of cases, that led to a second wave of COVID-19 in the greater areas of Seoul. As of 29 May, the number of cases that were linked to this cluster had reached 266 (KCDC, 2020a). Accordingly, the Seoul city government ordered all clubs, bars, and other nightlife establishments in the city to close indefinitely (KCDC, 2020a). Simultaneously, another cluster emerged from an e-commerce warehouse in the Gyeonggi Province, resulting in 108 cases as of 30 May.

In the last week of May, ∼40-80 daily new cases of COVID-19 were being reported (KCDC, 2020a). Following this spike in the number of new COVID-19 infections in nearly 2 months, public health authorities reimplemented strict lockdown measures in Seoul along with school closure, one more time across the nation. In June, it was announced that the strict social distancing campaign would be indefinitely extended as a preventive measure in Seoul, Incheon, and Gyeonggi Province; however, phased reopening of schools was initiated on May 20. It was reported by the KCDC that a holiday weekend in early May triggered a new wave of infections focused in the greater Seoul area, the so-called second wave of COVID-19 in South Korea (2020). In Seoul, the average number of new daily cases reported from 4 June to 17 June was 43 (KCDC, 2020a). This was followed by sporadic clusters of infections across the country in July, most of them associated with religious facilities and door-to-door salespeople, especially in the densely populated Seoul region and adjacent areas. Therefore, since 10 July, the government banned churches from organizing small gatherings other than regular worship services (KCDC, 2020a). As of 23 September, 23,216 cases of COVID-19 have been reported in South Korea, including 13.4% imported cases, 59.7% cases linked to local clusters, 14.5% unlinked local cases, and 12.4% cases under investigation (KCDC, 2020a).

To estimate the regional and temporal variability in the reproduction number of COVID-19 in South Korea, including the second wave concentrated in the greater Seoul areas, we analysed the spatiotemporal progression of the epidemic in the country from mid-February to mid-July 2020. Here our focus lies on estimating and interpreting the doubling time and effective reproduction number *R*_*t*_, a metric that quantifies the time-dependent transmission potential of the disease, incorporating the effect of control measures, susceptible depletion, and behavioural changes. This key epidemiological parameter, *R*_*t*_, represents the average number of secondary cases generated per case whenever conditions persist as they were at time *t*. Epidemic doubling times describes the sequence of intervals at which the cumulative incidence doubles (Lee et al., 2020, Muniz-Rodriguez et al., 2020). Therefore, an increase in the doubling time implies a decline in disease transmission. In this report, we estimated the doubling time and the effective reproduction number involving two epidemic waves of the COVID-19 epidemic in South Korea by employing the time series of cases by date of symptoms onset for the four most affected Korean regions: Seoul, Gyeonggi Province, Gyeongbuk Province, and Daegu. We also discuss the spatiotemporal variability of the reproduction number in terms of the public health policies that were put in place by the Korean government.

## Methods

### Data

We collected the daily series of confirmed local COVID-19 cases in South Korea from 20 January to 19 July, which were published by national and local public health authorities, including city or provincial departments of public health in South Korea (KCDC, 2020b). We focused our analysis on the regions with the highest caseloads including Seoul, Gyeonggi Province, Gyeongbuk Province, and Daegu (Figure 1).

**Figure 1.**
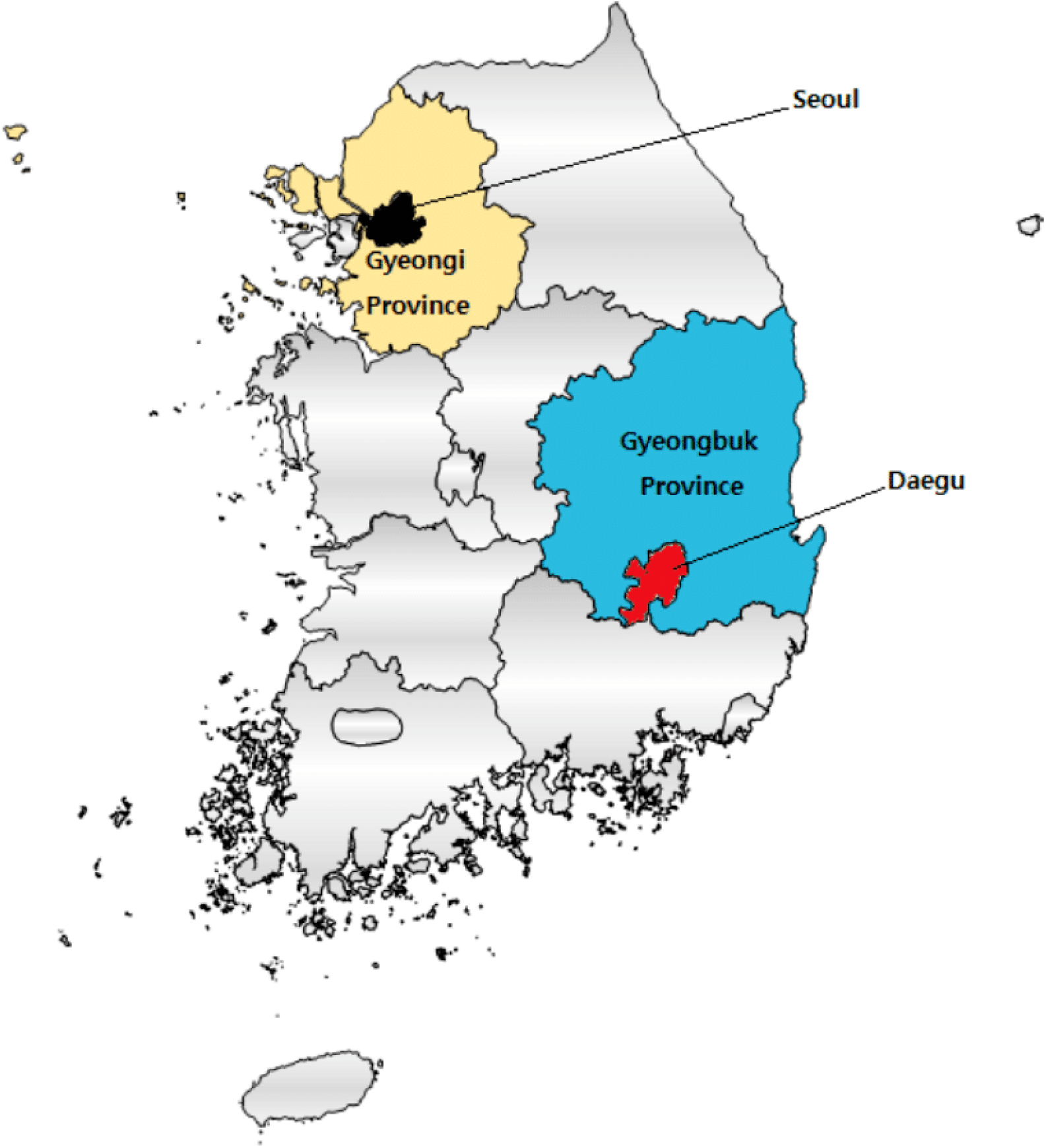
Map depicting the location of Seoul, Gyeonggi Province, Gyeongbuk Province, and Daegu.

### Imputing the date of onset

For a more accurate estimation of epidemic growth rates, the epidemic curve should be analyzed according to the date of symptom onset rather than the date of reporting because reporting delays can fluctuate substantially over the course of an epidemic. Reporting delays distort the incidence pattern of epidemics, misrepresenting the outbreak trajectory, thus possibly affecting the estimation of the reproduction number (Tariq et al., 2019). A prior study suggested that obtaining knowledge about reporting parameters, such as delay patterns and structure, improves the estimates of the reproduction numbers (Azmon et al., 2014). However, for the COVID-19 data in Korea, the date of symptom-onset is only available for 732 cases reported in Gyeonggi Province, which yielded a mean of 4.5 days and standard deviation of 4.4 days for the distribution of delays from symptoms onset to reporting of cases. Therefore, we utilized the empirical distribution of these 732 reporting delays from the onset of symptoms to reporting to impute the missing dates of onset for the remaining cases (Shim et al., 2020a). Specifically, we reconstructed 300 epidemic curves according to the date of symptom onset, from which we derived the mean incidence curve of local case incidence (Shim et al., 2020a, Tariq et al., 2019). For the calculation of *R*_*t*_(*t*), the mean incidence curve estimated based on the date of symptom onset was used for the regions of interest (i.e., Seoul, Gyeonggi Province, Gyeongbuk Province, and Daegu) (Figure 2). Using the reconstructed mean incidence curve of local case incidence, we removed the first and last three data points to adjust for the reporting delays in our real-time analysis. We assumed that the first wave ends when the mean incidence becomes less than 0.2 individuals per day. Similarly, we assumed that the second wave starts when the mean incidence of local cases becomes greater than 0.5 individuals per day. Slight variations to these thresholds did not affect our results.

**Figure 2.**
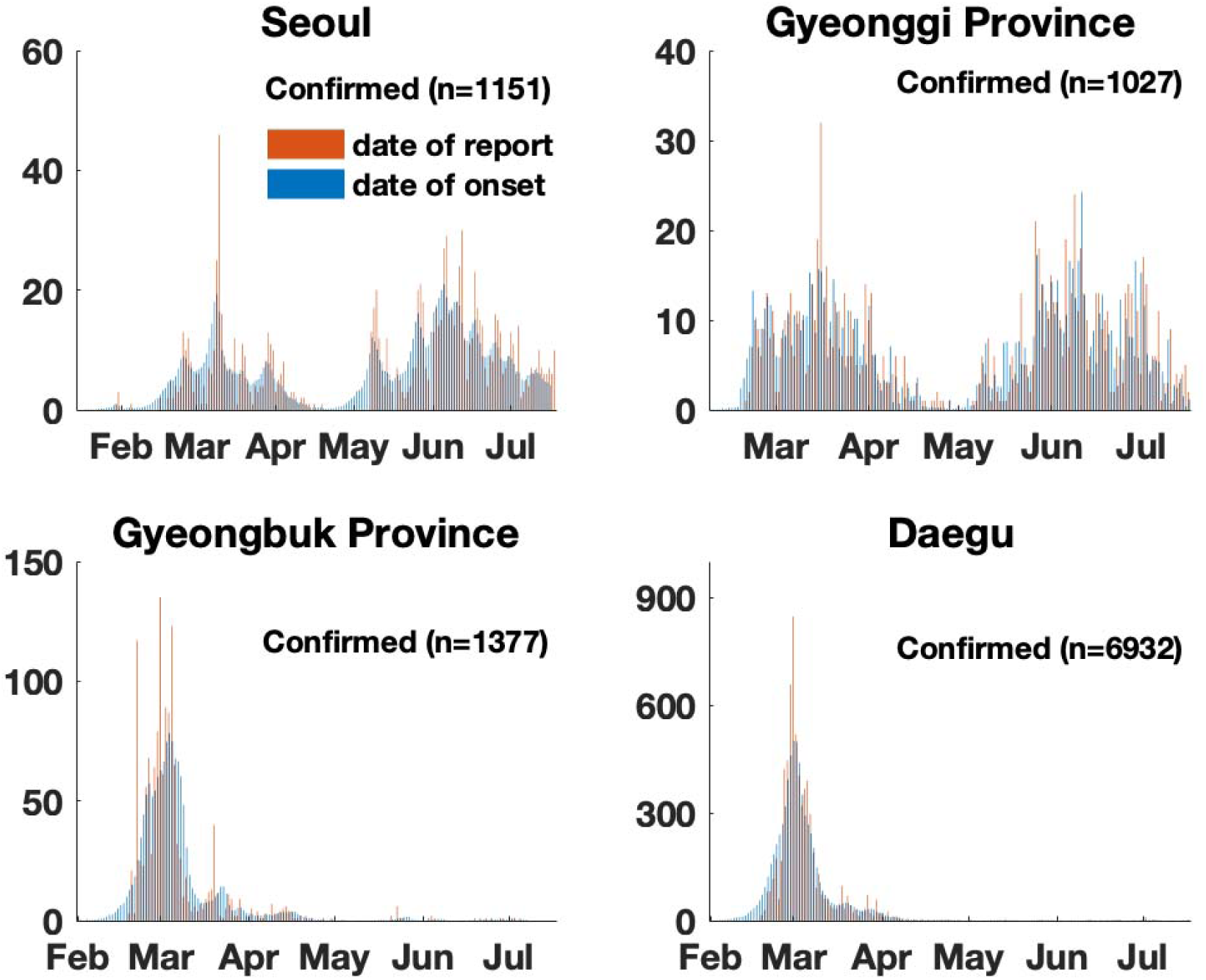
Timeline of confirmed cases of COVID-19 in Seoul, Gyeonggi Province, Gyeongbuk Province, and Daegu. The daily number of COVID-19 cases by date of report and by date of symptom onset are shown. The empirical distribution of reporting delays from the onset to diagnosis for 732 cases were used to impute the missing dates of onset for the remainder of the cases with missing data.

### Calculation of the doubling time

We analyzed the number of times COVID-19 cumulative incidence doubled and the evolution of the doubling times in the four most affected areas in Korea (i.e., Seoul, Gyeonggi Province, Gyeongbuk Province, and Daegu) from from 20 January to 19 July. Using regional-level daily cumulative incidence data, we calculated the times at which cumulative incidence doubles, denoted by 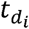 Specifically, we assume that

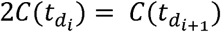

where 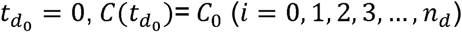, and 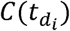 denotes the cumulative number of cases at time 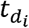 (Muniz-Rodriguez et al., 2020). Here, *n*_*d*_ is defined as the total number of times cumulative incidence doubles. Specifically, the sequence of doubling times are described as 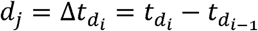 where *j* = 1, 2, 3, …, *n*_*d*_ In addition, we used parametric bootstrapping with a Poisson error structure around the harmonic mean of doubling times to obtain the 95% confidence interval (Chowell et al., 2006a, Chowell et al., 2006b).

### Calculation of R_t_

We assume that *R*_*t*_(*t*) can be estimated by the ratio of the number of new infections generated at time step *t* (*I*_*t*_) to the total infectiousness of infected individuals at time *t*, given by 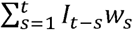 (Chong et al., 2018, Fraser, 2007a). Here, *w*_*s*_ denotes the infectivity profile of the infected individual, which is dependent on the time since infection (*s*) but independent of calendar time (*t*) (He et al., 2020, Wallinga and Teunis, 2004). Specifically, *w*_*s*_ is defined as a probability distribution describing the average infectiousness profile after infection. Individual biological factors such as pathogen shedding or symptom severity can affect the distribution *w*_*s*_. For example, an individual would be most infectious at time *s* when *w*_*s*_ is the largest. Thus 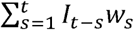 indicates the sum of infection incidence up to time step *t* − 1, weighted by the infectivity function *w*_*s*_. Steady values of *R*_*t*_ above one indicate sustained disease transmission, whereas values less than one indicate that the number of new cases is expected to follow a declining trend.

The infectivity profile, *w*_*s*_, can be approximated by the distribution of the generation time; however, times of infection are rarely observed, making it difficult to measure the distribution of the generation time (Fraser, 2007b). Therefore, the timing of symptoms onset is often used to estimate the distribution of the serial interval (SI) instead, which is defined as the time interval between symptom onset in two successive cases in a chain of transmission (Cori et al., 2013). Specifically, the infectiousness of a patient is a function of the time since infection and is proportional to *w*_*s*_ if we set the timing of infection in the primary case as the time zero of *w*_*s*_ and assume that the generation interval equals the SI. The SI was assumed to follow a gamma distribution with a mean of 4.8 days and a standard deviation of 2.3 days (Nishiura et al., 2020). Analytical estimates of *R*_*t*_ were obtained within a Bayesian framework using *EpiEstim R* package in *R* language version 3.6.3 (*R* Foundation for Statistical Computing, Vienna, Austria) (Cori et al., 2013). *R*_*t*_ was estimated at 7-day intervals, and we reported the median and 95% credible interval (CrI).

## Results

### City of Seoul

As of 19 July, Seoul has reported a total of 1,474 cases (10.7% of the total reported in South Korea), including 323 imported cases and 10 deaths, yielding an incidence rate estimated at 151 cases per million. In Seoul, the first peak based on the estimated dates of symptom onset occurred during the second week of March (8–14 March), with 18 new cases reported each day as the number of new cases linked to a Guro-gu call centre kept rising. Based on the estimated dates of symptom onset, the 7-day moving average of daily cases reached 19 cases on 9 March (Figure 3) whereas the highest value of *R*_*t*_ was estimated at, *R*_*t*_ ∼2.9 (95% CrI: 1.6-4.7) on 19 February which continued to stay above one until 6 March (Figure 3).

**Figure 3.**
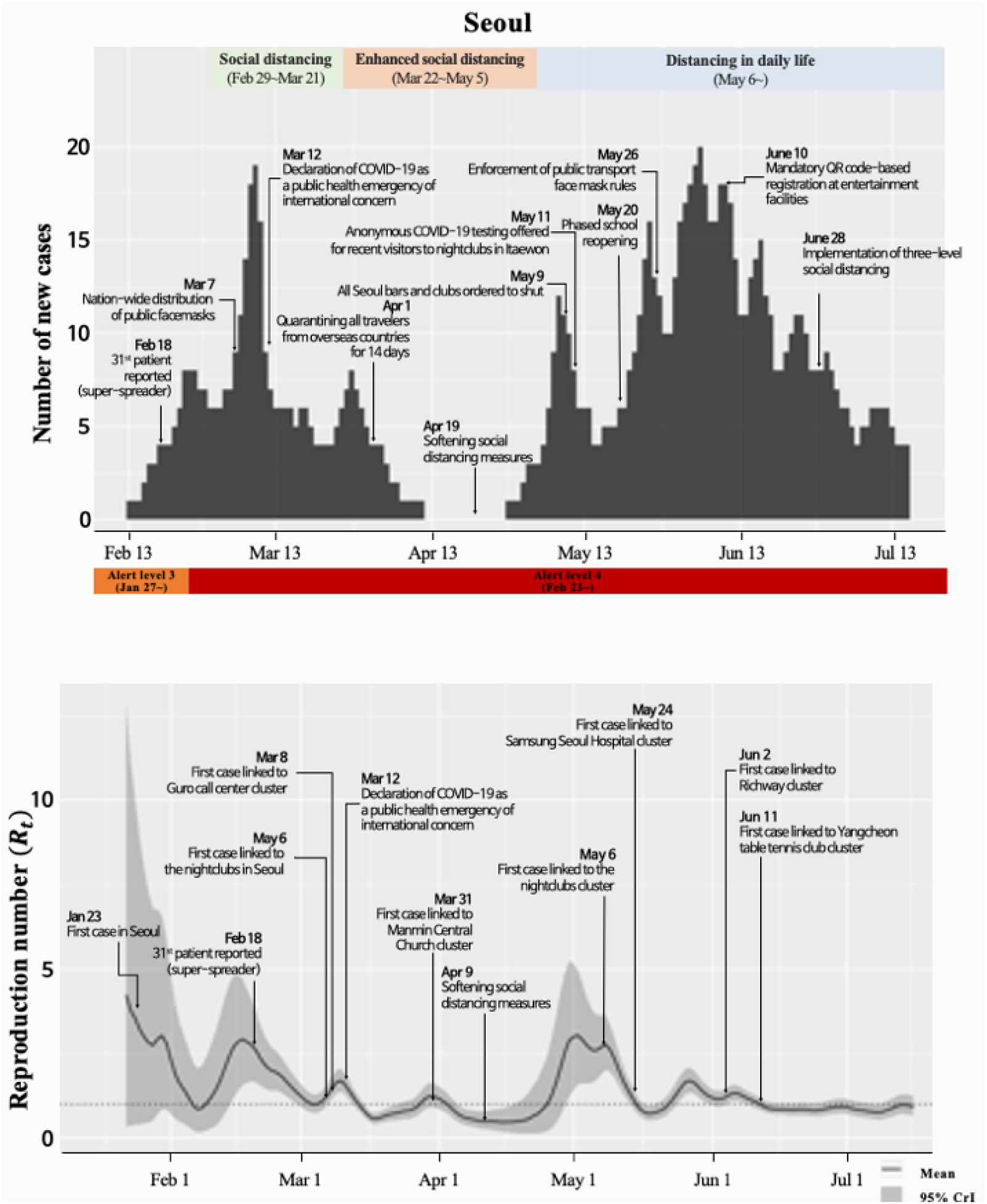
The epidemic trajectory of coronavirus disease 2019 in Seoul, as of 19 July 2020. Upper panel: The epidemic curve shows the daily number of new cases by the imputed date of symptom onset. The date of symptom onset for cases with missing data was imputed based on the empirical distribution of the delay from the onset of symptoms to reporting. Lower panel: Real-time estimates of the time-varying reproduction number (*R*_*t*_) in Seoul. The solid line indicates the daily estimated *R*_*t*_ and the grey area indicates the 95% credible interval of *R*_*t*_. The dotted line indicates the epidemic threshold of *R*_*t*_ =1.

After its first peak in February, the number of daily new cases by date of symptoms onset in Seoul gradually declined, dropping below five on 1 April and staying under five new cases per day for about a month (Figure 3). However, in early May, despite a steady decline in imported cases, locally transmitted infections surged throughout the Seoul metropolitan area with case clusters traced to clubs, churches, and sports facilities. Therefore, *R*_*t*_ increased, reaching 3.0 (95% CrI: 1.6-5.0) on 4 May 2020. During the first wave, the doubling time was estimated to be 7.5 (95% CI: 7.0, 8.2) days in Seoul (Table 1).

**Table 1.** Regional variations in doubling times in days of COVID-19 cumulative incidence and its 95% CI, from 20 January to 19 July, 2020: Seoul, Gyeonggi Province, Gyeongbuk Province, and Daegu.

| Regions | Mean doubling time (95% CI) |  |
| --- | --- | --- |
|  | First wave | Second wave |
| Seoul | 7.5 (7.0, 8.2) | 6.0 (5.4, 6.7) |
| Gyeonggi Province | 4.6 (4.2, 5.5) | 7.5 (5.4, 8.6) |
| Gyeongbuk Province | 3.6 (3.5, 4.0) | 10.1 (4.6,14.5) |
| Daegu | 2.8 (2.5, 4.0) | 10.0 (7.1, 13.4) |

The number of cases continued to increase thereon, and in the first week of June, the average daily number of confirmed COVID-19 cases in the capital surpassed the previous high point in the daily number of confirmed cases in the middle of March. The major clusters in Seoul were linked to nightclubs (139 cases), the Guro-gu call centre (99 cases), Manmin Central Church (41 cases), Richway (97 cases), Yangcheon-gu table tennis club (41 cases), and Newly Planted Church in the Seoul Metropolitan Region (37 cases) as of 18 June. On 14 June, the average *R*_*t*_ in the capital, which reflects the average number of people infected by a patient, dropped below one (95% CrI: 0.8-1.2), implying that the spread of the virus had slowed down substantially in the city (Figure 3). During the second wave, the doubling time in Seoul decreased to 6.0 (95% CI: 5.4, 6.7) days, indicating faster transmission compared to that during the first wave (Table 1). As of 15 July 2020, the *R*_*t*_ in Seoul was estimated at 0.9 (95% CrI: 0.7-1.2), straddling the epidemic threshold of 1.0, and suggesting potential for further transmission of the virus.

### Gyeonggi Province

Gyeonggi Province (literally meaning the “province surrounding Seoul”) is located in the western central region of Korea and is the most populous province in South Korea with a population of 13.5 million people. In Gyeonggi Province, the daily number of new cases by date of symptoms onset during the last weeks of February was 6.3 on average (Figure 4). Accordingly, the first peak of *R*_*t*_ occurred on 22 February, reaching 8.9 (95% CrI: 4.8-14.2), with an estimated doubling time of 4.6 (95% CI: 4.2, 5.5) days (Table 1). In the second week of March, South Korea recorded continuous drops in the number of daily new infections as massive testing of the followers of a religious sect in the south-eastern city of Daegu, the epicentre of COVID-19, was nearing its end; thereafter, the number of cases in Gyeonggi Province gradually decreased.

**Figure 4.**
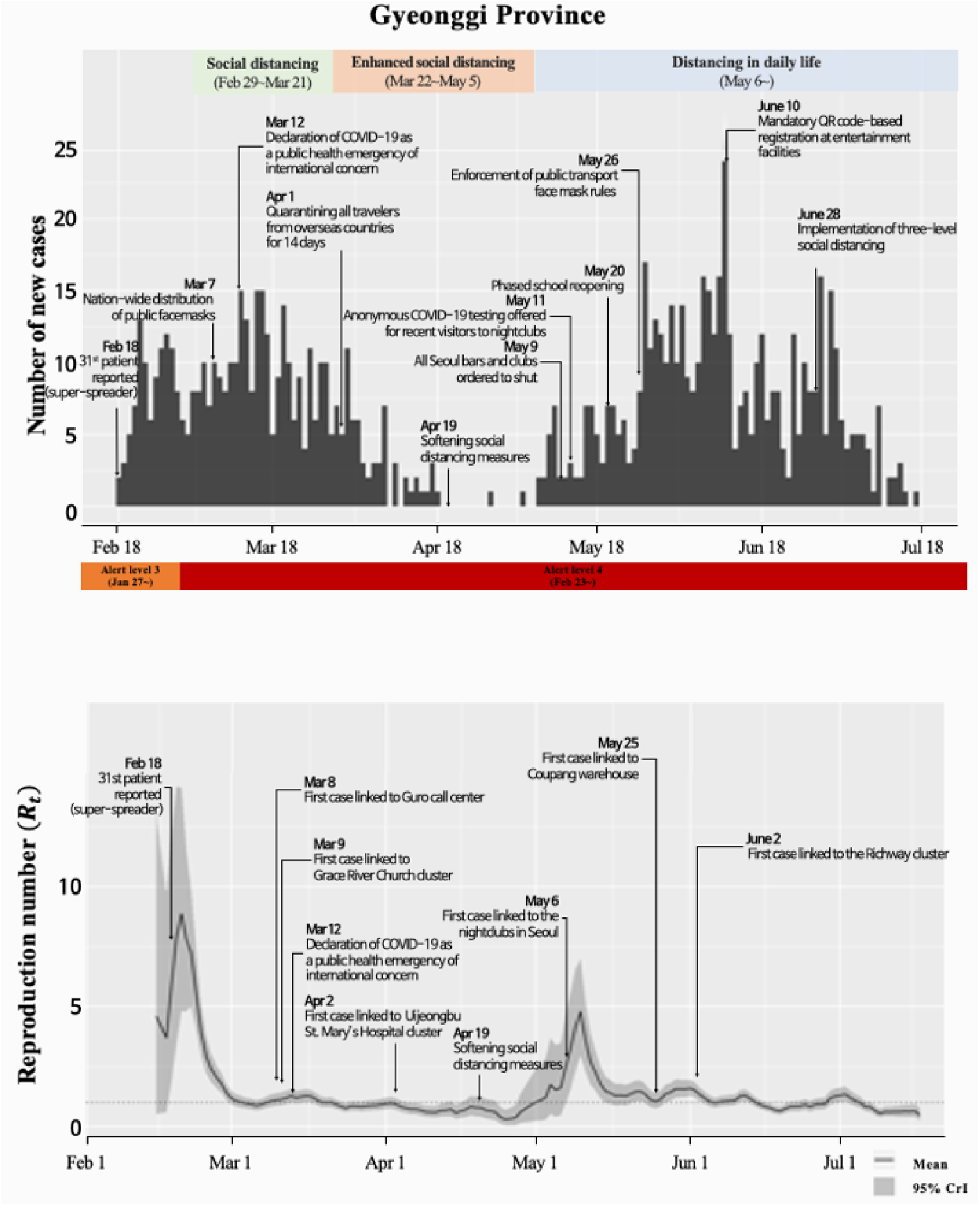
The epidemic trajectory of coronavirus disease 2019 in Gyeonggi Province, as of 19 July 2020. Upper panel: The epidemic curve shows the daily number of new cases by the imputed date of symptom onset. The date of symptom onset for cases with missing data was imputed based on the empirical distribution of the delay from the onset of symptoms to reporting. Lower panel: Real-time estimates of the time-varying reproduction number (*R*_*t*_) in Gyeonggi Province. The solid line indicates the daily estimated *R*_*t*_ and the grey area indicates the 95% credible interval of *R*_*t*_. The dotted line indicates the epidemic threshold of *R*_*t*_ =1.

However, clusters of infections in Gyeonggi Province raised concerns about further community spread, with a resurgence of cases in the province occurring in late May and resulting in the highest peak in early June. Between 1-13 June, an average of 14 new cases were reported each day in Gyeonggi Province. The second peak of *R*_*t*_ in the region occurred on 12 May, with an estimated value of *R*_*t*_ at 4.8 (95% CrI: 3.0-7.0) and the doubling time estimated at 7.5 (95% CI: 5.4, 8.6) days (Table 1). Since its second peak, *R*_*t*_ gradually decreased (Figure 4); however, a series of sporadic clusters have continued to occur. The major clusters in Gyeonggi Province included Grace River Church (67), Coupang warehouse (67), nightclubs (59), Richway (58), Uijeongbu St. Mary’s Hospital (50), Guro-gu call centre/Bucheon SaengMyeongSu Church (50), door-to-door sales in the Seoul Metropolitan Region (32), and Yangcheon-gu sports facility (28). As of 19 July, the number of local cases in Gyeonggi Province was 1,027 (10.4% of the total reported cases in South Korea), including 29 deaths, with an *R*_*t*_ estimated at 0.8 (Figure 4). The incidence rate in the province was estimated at 108 per million.

### Gyeongbuk Province

The first case in the Sincheonji cult cluster (the largest COVID-19 cluster in South Korea) appeared on 18 February, resulting in sustained transmission chains, with 39% of the cases associated with the church cluster in Gyeongbuk Province. Therefore, the virus alert level was raised to “red” (the highest level) on 23 February, and health authorities focused on halting the spread of the virus in Daegu and Gyeongbuk Provinces. Figure 5 shows that the peak of the epidemic occurred in the first week of March (with a reproduction number greater than one until 9 March) (Figure 5). The doubling time in Gyeongbuk Province reached the values as short as 3.6 (96% CI: 3.5, 4.0) days (Table 1). As of 18 July, the number of cases in Gyeongbuk Province was 1,393, including 54 deaths. Among these cases, 566 were related to the Shincheonji cluster. The incidence rate in Gyeongbuk Province was 523 per million, accounting for 10.2% of all confirmed cases in South Korea (KCDC, 2020a). The major clusters in Gyeongbuk Province were linked to Cheongdo Daenam Hospital (119 cases), Bonghwa Pureun Nursing Home (68 cases), Gyeongsan Seo Convalescent Hospital (66 cases), pilgrimage to Israel (41 cases), Yecheon-gun (40 cases), and Gumi Elim Church (11 cases).

**Figure 5.**
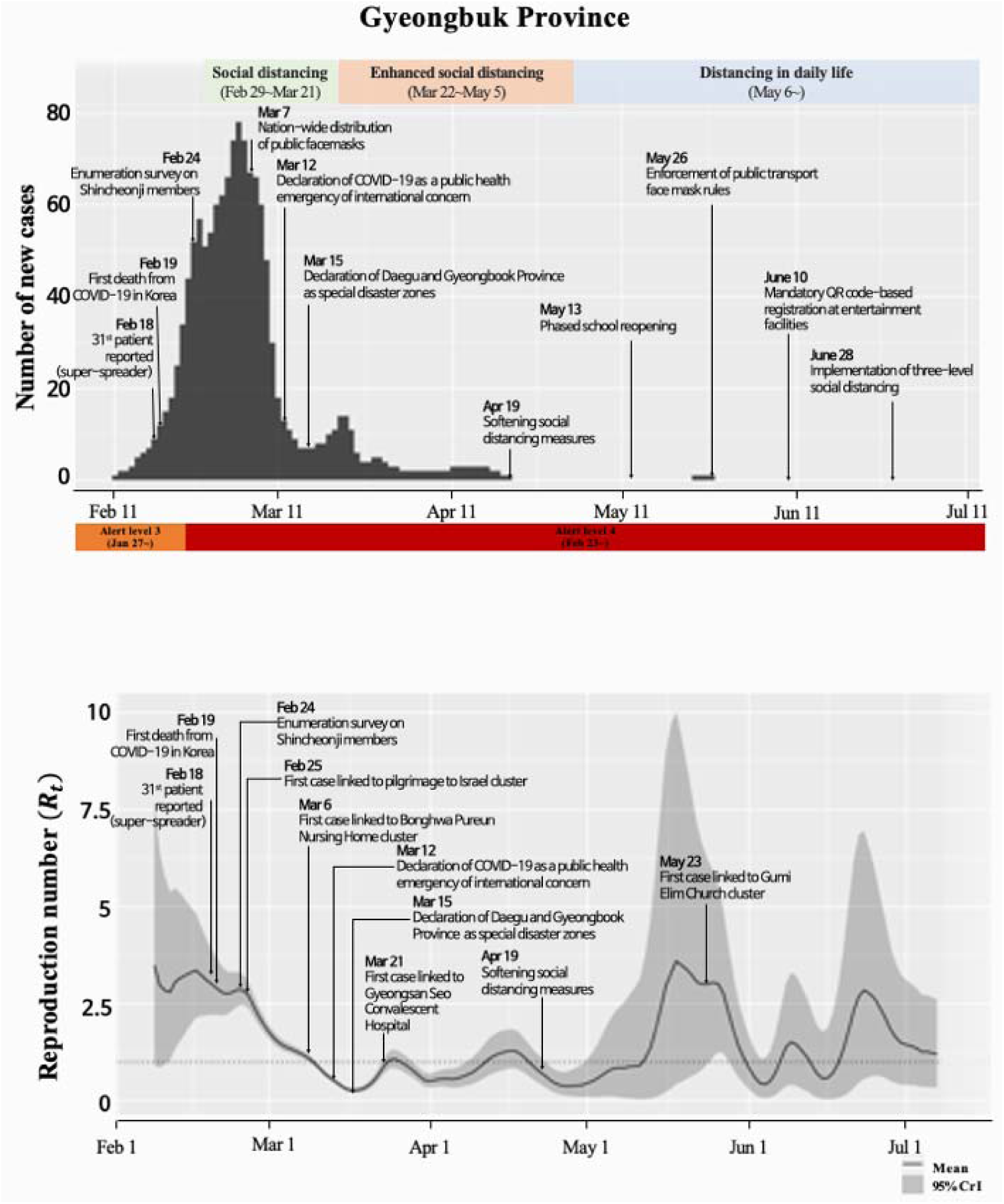
The epidemic trajectory of coronavirus disease 2019 in Gyeongbuk Province, as of 19 July 2020. Upper panel: The epidemic curve shows the daily number of new cases by the imputed date of symptom onset. The date of symptom onset for cases with missing data was imputed based on the empirical distribution of the delay from the onset of symptoms to reporting. Lower panel: Real-time estimates of the time-varying reproduction number (*R*_*t*_) in Gyeongbuk Province. The solid line indicates the daily estimated *R*_*t*_ and the grey area indicates the 95% credible interval of *R*_*t*_. The dotted line indicates the epidemic threshold of *R*_*t*_ =1.

### City of Daegu

The epicentre of the South Korean COVID-19 outbreak has been identified in Daegu, a city of 2.5 million people, approximately 150 miles south-east of Seoul. The rapid spread of COVID-19 in Daegu was attributed to a superspreading event in a religious group called Shincheonji, resulting in an explosive outbreak with 4,511 infections in the city of Daegu, resulting in the relatively short doubling time, i.e. 2.8 (95% CI: 2.5, 4.0) days (Table 1 and Figure 6). Other major clusters in Daegu included the second Mi-Ju Hospital (196 cases), Hansarang Convalescent Hospital (124 cases), Daesil Convalescent Hospital (101 cases), and Fatima Hospital (39 cases). Daegu was the most severely affected area in South Korea with 6,932 cumulative cases as of 19 July, accounting for 51.0% of all confirmed cases in Korea. According to our model, the number of new cases based on the onset of symptoms was estimated to be the highest on 27 February; the number gradually decreased thereafter. Accordingly, the estimated Rt was above two until 27 February and dropped below one on 5 March, although recent sporadic infections caused the *R*_*t*_ to fluctuate around one (Figure 6).

**Figure 6.**
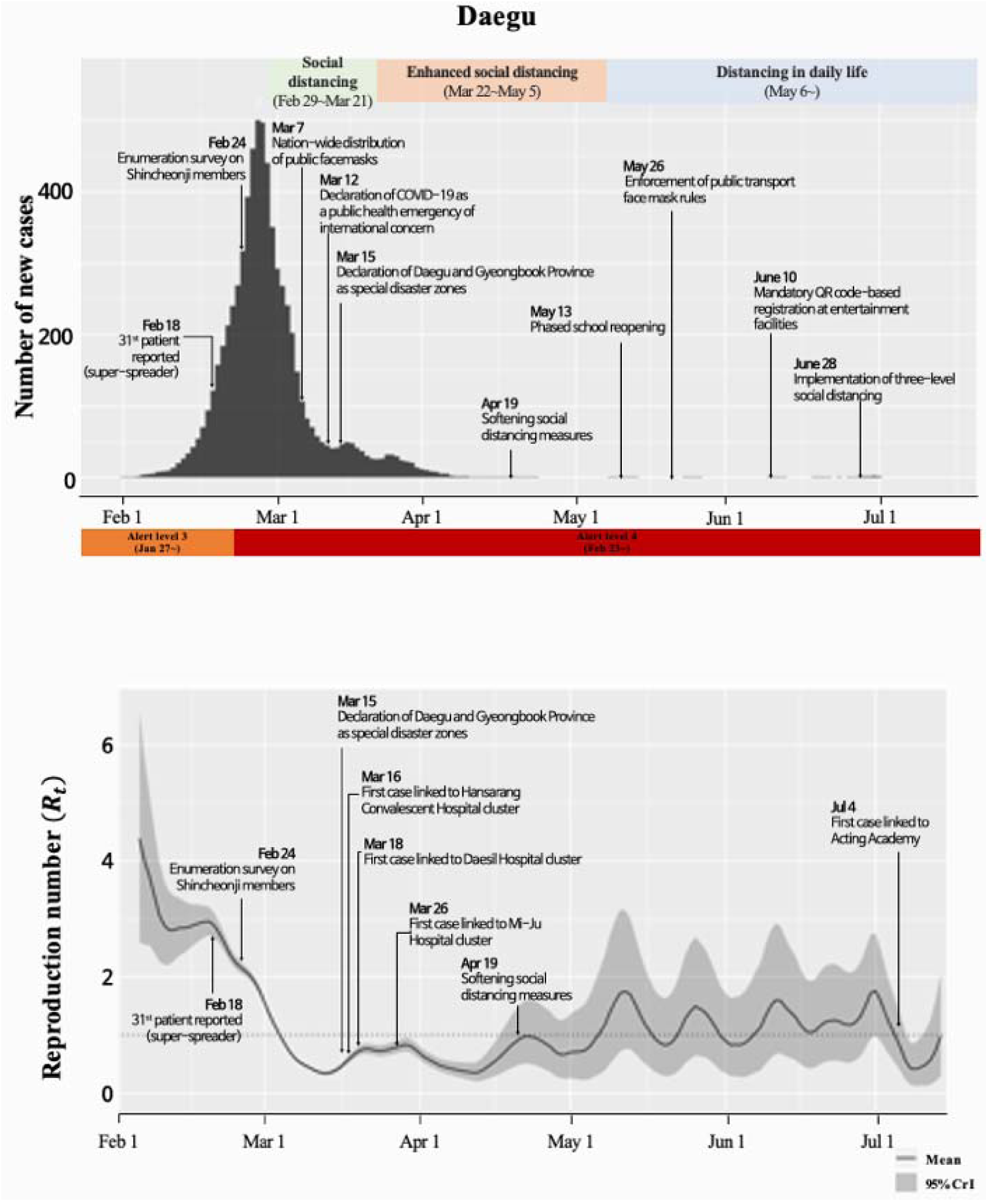
The epidemic trajectory of coronavirus disease 2019 in Daegu, as of 19 July 2020. Upper panel: The epidemic curve shows the daily number of new cases by the imputed date of symptom onset. The date of symptom onset for cases with missing data was imputed based on the empirical distribution of the delay from the onset of symptoms to reporting. Lower panel: Real-time estimates of the time-varying reproduction number (*R*_*t*_) in Daegu. The solid line indicates the daily estimated *R*_*t*_ and the grey area indicates the 95% credible interval of *R*_*t*_. The dotted line indicates the epidemic threshold of *R*_*t*_ =1.

## Discussion

The estimates of the transmission potential of COVID-19 in Korea displayed substantial spatiotemporal variation. Indeed, several factors influence the value of the reproduction number, including the transmissibility of an infectious agent, individual susceptibility, individual contact rates, and control measures (Anderson and May, 1991). Our results indicate that the effective reproduction number for COVID-19 declined to low levels after the first wave and straddled around the epidemic threshold of 1.0 in March and April suggesting that social distancing measures had a significant effect on mitigating the spread of the novel coronavirus. Estimates of early national *R*_*t*_ for South Korea retrieved from other studies, 2.9 (95% CrI 2.0-3.9) in February (Ryu et al., 2020) and 2.6 (95% CI: 2.3-2.9) in March, are in good agreement with our *R*_*t*_ estimates (Zhuang et al., 2020).

Our results suggest that South Korea has experienced two spatially heterogenous waves of the novel coronavirus. At the regional level, Seoul and Gyeonggi Province have experienced two waves whereas Daegu and Gyeongbuk Provinces are yet to experience the second wave of the disease. The highest epidemic peak occurred in Daegu and Gyeongbuk Province in late February and early March, with *R*_*t*_ estimated at 4.4 (95% CrI: 2.6-6.6) and 3.5 (95% CrI: 0.9-7.3), respectively. During their epidemic peak, the doubling time was estimated at 2.8 (95% CI 2.5, 4.0) days and 3.6 (95% CI 3.5, 4.0) days in Daegu and Gyeongbuk Province, respectively, which is similar to prior estimates of doubling time, 3.8 (95% CI: 3.4-4.2) days (Lee et al., 2020). Similarly, in Gyeonggi Province and Seoul, the first wave was observed in late February and early March, respectively. However, sporadic clusters of infections appeared in Seoul and near Gyeonggi Province, immediately after the government eased its strict nationwide social distancing guidelines on May 6. This resurgence of infections in Seoul and Gyeonggi Province (i.e., the province surrounding Seoul) after a sustained period with fewer than 5 cases per day in each region, led to the second epidemic wave with sub-exponential growth dynamics. In Seoul, the mean doubling time decreased from 7.5 (95% CI: 7.0, 8.2) days during the first wave to 6.0 (95% CI: 5.4, 6.7) days during the second wave, indicating faster transmission during the case resurgences. Accordingly, our findings revealed sustained local transmission in Seoul and Gyeonggi Province, with the estimated reproduction number estimated above one until the end of May. In late May, the country implemented two weeks of strict social distancing measures incorporating stringent virus prevention guidelines for the metropolitan area. These measures included the shutting down of public facilities and regulating bars and karaoke rooms. In the second week of June, South Korea decided to indefinitely extend a period of strict social distancing measures, as nearly all locally transmitted cases were in the metropolitan area.

Although Korea has a relatively low number of reported cases compared to other countries including the U.S. and China, it is believed that South Korea is currently experiencing yet another resurgence of the virus (WHO). Originally, South Korean authorities predicted a resurgence of the virus in the fall or winter; however, this possible second wave started in and around Seoul, which, with 51.6 million inhabitants, accounts for about half of the entire population of the country. Secondary waves of the disease can result from multiple factors, including easing of travel restrictions and resuming social activities especially in the high population density areas of Seoul and Gyeonggi Province. Furthermore, a substantial proportion of COVID-19 cases are asymptomatic (Mizumoto et al., 2020); thus, they are not detected by surveillance systems, resulting in the underestimation of the epidemic growth curve. It was also recently reported that individuals aged 20-39 years in South Korea drove the COVID-19 epidemic throughout society with multiple rebounds, and an increase in infection among the elderly was significantly associated with an elevated transmission risk among young adults (Yu et al., 2020).

Our study is not exempt of limitations including the lack of dates of symptoms onset for all of the cases, relying on a statistical reconstruction of the epidemic curve by dates of symptoms onset as in a previous study (Shim et al., 2020a). Overall, using most up-to-date epidemiological data from South Korea, our study highlights the effectiveness of strong control interventions in South Korea and emphasizes the need to maintain firm social distancing and contact tracing efforts to mitigate the risk of additional waves of the disease.

## Data Availability

www.kcdc.co.kr

## Contributions

ES conceptualized analysis, retrieved and managed the data. ES, GC, and AT analyzed the data. ES and GC wrote the first draft of the paper. All authors contributed to the writing of the paper.

## Ethical Approval

Not required.

